# Mobile Stroke Unit Treatment Times and Transport Frequency in a Suburban Setting

**DOI:** 10.1101/2023.03.02.23286726

**Authors:** Alejandro Vargas, Rebecca Zobel, Laurel J. Cherian, Rima M. Dafer, Nicholas D. Osteraas, Sarah Y. Song, Mark E. Cichon, James J. Conners

## Abstract

**Background:** The majority of Mobile Stoke Units (MSUs) operate in European and United States urban cities. Questions remain on the cost-effectiveness, setting (urban, suburban, or rural), infrastructure and support, and reimbursement of these units. We present our experiences of a single-center MSU in a suburban setting, with treatment times, challenges, and possible future directions of alternative methods of care.

**Methods:** Retrospective analysis of prospectively collected data from Mobile Stroke Unit calls for service and Get With The Guidelines-Stroke data from two primary stroke centers from December 2017 through February 2020 comparing patients receiving intravenous thrombolysis and treatment times.

**Results:** There were no differences in age, sex, medical history, or stroke severity between MSU transport when compared to standard transport. There were differences in patient racial and ethnic demographics between groups, with higher white race and Hispanic ethnicity. Door-to-needle time was 48.9 minutes for patients seen on the Rush MSU versus 67.2 minutes for patients seen via traditional EMS transport (p=0.04).

**Conclusions:** The Rush MSU demonstrated significant reduction of acute ischemic stroke treatment time with intravenous thrombolysis, but did not demonstrate the patient volume necessary to justify continued operation. Suburban and rural regions do benefit from pre-hospital stroke evaluation, however the ideal method for a cost-effective strategy is still unknown.

## Introduction

Mobile Stroke Units (MSUs) were first conceived in 2003 as a means to rescue brain tissue from ischemic damage through faster acute thrombolysis treatment, thus reducing individual suffering and life-long cost. It has been postulated that MSU-related costs may be outweighed by possible reduction of disability [1]. Reduction in treatment times with thrombolytics with various MSUs have shown improvement in functional outcomes [2,3]. Since the majority of MSUs operate in European and United States urban cities, questions remain on the cost-effectiveness, setting (urban, suburban, or rural), infrastructure and support, and reimbursement of these units [4]. We present our experiences of a single-center MSU in a suburban setting, with treatment times, challenges, and possible future directions of alternative methods of care.

## Methods

### Study Design

This study was a retrospective analysis of prospectively collected data from MSU calls for service and Get With The Guidelines-Stroke data from December 2017 through February 2020. We compared MSU patients to patients transported by traditional EMS methods to two primary stroke centers in Oak Park, IL, a suburb of Chicago, Illinois with a population of 53,224. One of these centers served as the home base of the Rush MSU. The same vascular neurologists providing telestroke consultation on the MSU saw the patients via telestroke in the emergency departments of both primary stroke centers if transported by traditional EMS methods. The study protocol was approved by our Institutional Review Board.

### Mobile Stroke Unit Standard Operating Procedures

The Rush MSU operated 12 hours a day (7AM to 7PM), 7 days a week from December 2017 until termination of the program in February 2020. There was a brief shift to 24-hour coverage between October 2018 to May 2019, until return to 12 hours was favored for resource allocation. Dispatch was simultaneous along with local EMS unit for all 911 calls identified as suspected stroke. The MSU was staffed by a specially trained registered nurse (RN), CT technologist, paramedic, and emergency medical technician. If local EMS arrived prior to the MSU and there was clinical suspicion for a non-stroke diagnosis, then local EMS remained primary and the MSU left the scene. If at any time, airway, breathing, or circulation was compromised, these issues took precedence in accordance to regional EMS protocols and standard operating procedures.

Patients with focal neurological deficits <24 hours old were accepted onto the MSU if first contact on scene or co-arrival with local EMS; this was determined as “door” time. A head CT was immediately obtained for all patients. Once the patient completed the head CT, the vascular neurologist remotely performed a National Institutes of Health Stroke Scale (NIHSS) via live two-way audio-video communication and any imaging was simultaneously reviewed by the vascular neurologist and reading neuroradiologist (at the home base hospital) to determine intravenous tissue plasminogen activator (tPA) eligibility. Patients with signs and symptoms of a large vessel occlusion with symptom onset <24 hours could have CT angiography (CTA) obtained and transported to the closest of the two regional comprehensive stroke centers. The first center was Loyola University Medical Center (a 547 bed academic medical center 2.9 miles from the Rush MSU base), and the second center was Rush University Medical Center (a 671 bed academic medical center 7.7 miles from the Rush MSU base). In these cases, the MSU provided pre-arrival notification to the emergency department and neuro-interventional team to facilitate advanced preparation for possible emergency treatment.

### Statistical Analysis

We summarized all collected information for both groups using descriptive statistics, including mean, median, and standard deviation. We evaluated the differences in baseline characteristics and treatment time metrics between groups. Continuous variables were reported as mean and standard deviation (SD) and were compared with the student’s t or Mann-Whitney tests. Categorical variables were reported as proportions and compared with the χ2 and Fisher’s exact test. The analysis was performed using SPSS Version 22.0, and the level of significance was established at a 0.05 level (2-sided). Door time for MSU was from when the patient entered the MSU while door-to-needle (DTN) was reported as usual care for traditional EMS transport.

## Results

The Rush MSU service area at the time of closure included 9 separate towns or villages in suburban Cook County, within the Chicago metropolitan area, and a maximum population of 207,801. During the 27-month period of December 2017 to February 2020 when the MSU was in service, 266 dispatches were performed, and 135 (50.8%) of these were patient transports. This was an average of 9.9 dispatches, or 5 patient transports a month. IV tPA was administered 14 times during these 27 months, with only one being administered outside of the 7AM to 7PM time, with an average of one IV tPA administration every two months. During the same time frame, 86 patients received IV tPA in two surrounding primary stroke centers, with an average of 3.2 IV tPA administrations per month.

Patient demographics were self-reported when prospectively collected. Mean patient age was 70.1 years (SD 12.0) on the Rush MSU versus 65.8 years (SD 15.4) via traditional EMS transport (p=0.42). Male sex was recorded in 6 patients (42.9%) on the Rush MSU versus 43 (50.0%) via traditional EMS transport (p=0.78). Race was significantly different between groups, with Rush MSU patients 78.6% white and 14.3% black, while traditional EMS transport patients were 37.2% white and 60.5% black (p=0.01). Hispanic ethnicity was also significantly different between both groups, with 4 patients of Hispanic ethnicity (28.6%) on the Rush MSU versus 5 patients (5.9%) via traditional EMS transport (p-value 0.02). There were no significant baseline differences in self-reported history of hypertension, diabetes mellitus, hyperlipidemia, coronary artery disease, previous stroke, or atrial fibrillation were seen. The demographics and baseline characteristics are summarized in table 1.

**Table 1:** Patient demographics, risk factors, stroke severity, and treatment times among patients receiving intravenous alteplase on the Rush Mobile Stroke Unit (MSU) compared to traditional Emergency Medical Services (EMS) transport

| Variable | MSU (n=14) | EMS (n=86) | p-value |
| --- | --- | --- | --- |
| Age (years), mean (SD) | 70.1 (12.0) | 65.8 (15.4) | 0.42 |
| Male sex, n (%) | 6 (42.9%) | 43 (50.0%) | 0.78 |
| Race, n (%): |  |  | **0.01** |
| White | 11 (78.6%) | 32 (37.2%) |  |
| Black | 2 (14.3%) | 52 (60.5%) |  |
| Asian | 0 (0.0%) | 2 (2.3%) |  |
| American Indian / Alaskan Native | 1 (7.1%) | 0 (0.0%) |  |
| Hispanic ethnicity, n (%) | 4 (28.6%) | 5 (5.9%) | **0.02** |
| Hypertension, n (%) | 10 (71.4%) | 51 (59.3%) | 0.56 |
| Diabetes Mellitus, n (%) | 6 (42.9%) | 16 (18.6%) | 0.08 |
| Hyperlipidemia, n (%) | 4 (28.6%) | 16 (18.6%) | 0.47 |
| Coronary Artery Disease, n (%) | 1 (7.1%) | 11 (12.8%) | 1.00 |
| Previous Stroke, n (%) | 6 (42.9%) | 15 (17.4%) | 0.07 |
| Atrial Fibrillation, n (%) | 0 (0.0%) | 10 (11.6%) | 0.35 |
| Initial NIHSS (score), mean (SD) | 10.1 (10.2) | 10.2 (7.9) | 0.96 |
| DTN (minutes), mean (SD) | 48.9 (7.8) | 67.2 (29.6) | **0.04** |
| OTN (minutes), mean (SD) | 112.9 (60.3) | 133 (60.3) | 0.17 |

Key: Mobile Stroke Unit (MSU), Emergency Medical Services (EMS), standard deviation (SD), National Institutes of Health Stroke Scale (NIHSS), Door-to-needle (DTN), Onset to needle (OTN)

There was no difference in stroke severity per NIHSS score, with mean NIHSS 10.1 (SD 10.2) for Rush MSU, and 10.2 (SD 7.9) for traditional EMS transport (p=0.96). When considering treatment times, DTN time was 48.9 minutes (SD 7.8) for patients seen on the Rush MSU versus 67.2 minutes (SD 29.6) for patients seen via traditional EMS transport (p=0.04).

Onset to needle (OTN) time was 112.9 minutes (SD 60.3) for patients seen on the Rush MSU versus 133 minutes (SD 60.3) for patients seen via traditional EMS transport (p=0.17). 2 of the 14 patients (14.3%) were treated under 60 minutes from stroke onset on the Rush MSU versus 8 of 81 (9.9%) with 5 missing times for patient seen via traditional EMS transport.

## Discussion

Our single-center MSU demonstrated significant reduction of treatment with intravenous tPA in comparison to two primary stroke centers in a suburban setting. Currently, the majority of MSUs operate and serve in urban metropolitan areas except for the University of Homburg / Saarland, and the University of Alberta, with limited data for MSUs in suburban and rural settings [5,6]. The high cost of MSUs may limit widespread use in all population-density models. One MSU may cost 1 million USD to start, and 1 million USD to maintain per year, and would need sufficient volume to justify cost [4].

Ideal methods of MSU protocols in more remote settings are yet to be determined, with rendezvous coordination or Air MSUs as possible strategies [7]. Prioritizing rapid evaluation or rapid imaging in more remote areas may also guide infrastructure and funding decisions for pre-hospital evaluations. Using existing tele-stroke or tele-neurology networks for pre-hospital evaluation of patients, such as tablet-based evaluations within existing ambulance infrastructure, may provide more accurate triage and pragmatic resource utilization in remote settings [8]. There also needs to be sufficient buy-in by local EMS networks and leaders to maximize patient triage and treatment. Our MSU at the time of closure did not have complete coverage of the proposed region and was only able to provide service to less than half of the proposed population (Figure 1). There were also substantially more patients receiving thrombolysis in the ED than in the MSU, which may reflect any of the steps involved, including 911 training and triage, local EMS buy-in to the program, familiarity with protocol, and living in coverage area of the MSU. It is an interesting finding that the racial and ethnic make-up was significantly different between ED and MSU patients.

**Figure 1:**
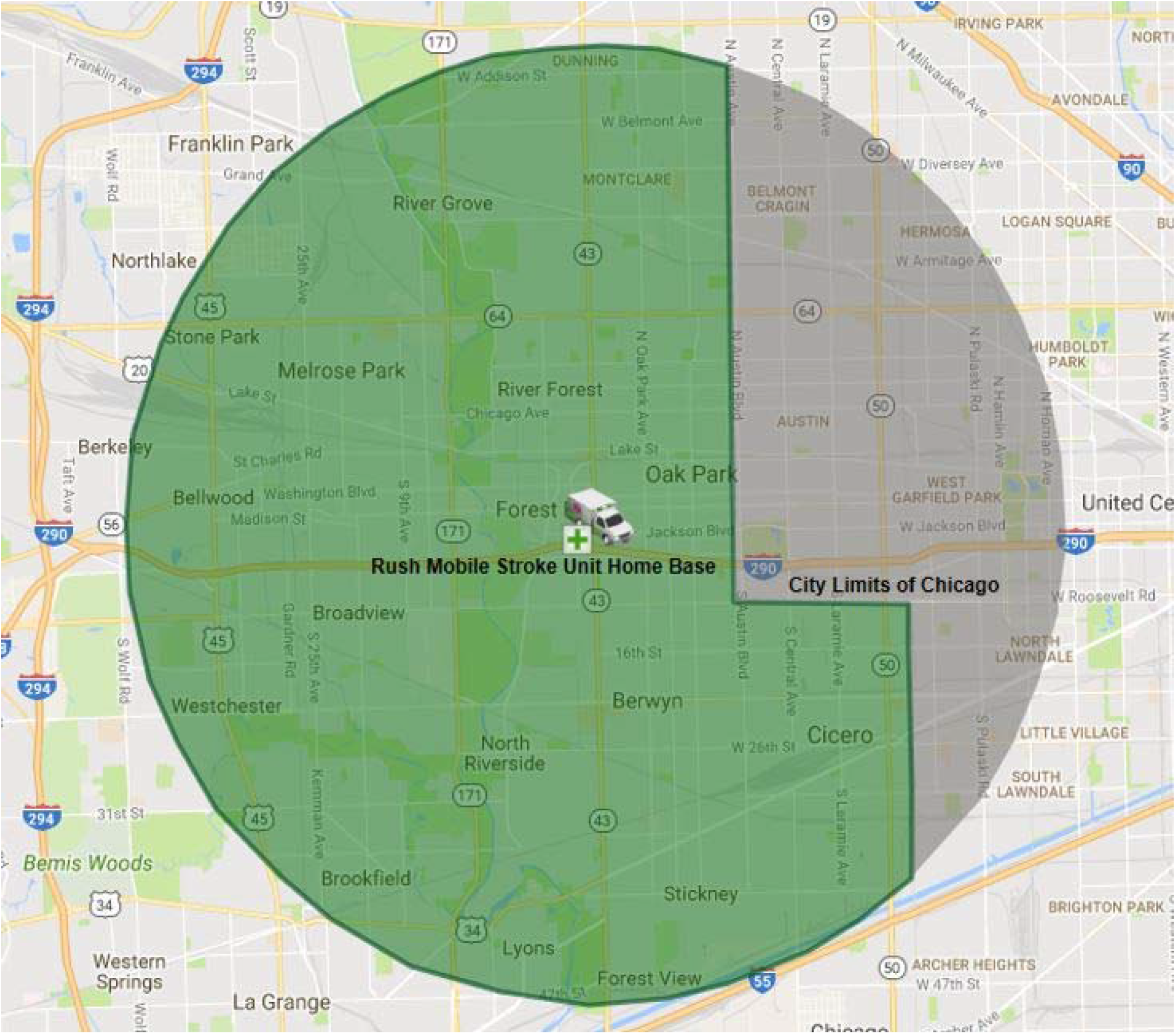
Initial proposed 5-mile radius coverage of the Rush Mobile Stroke Unit

**Figure 2:**
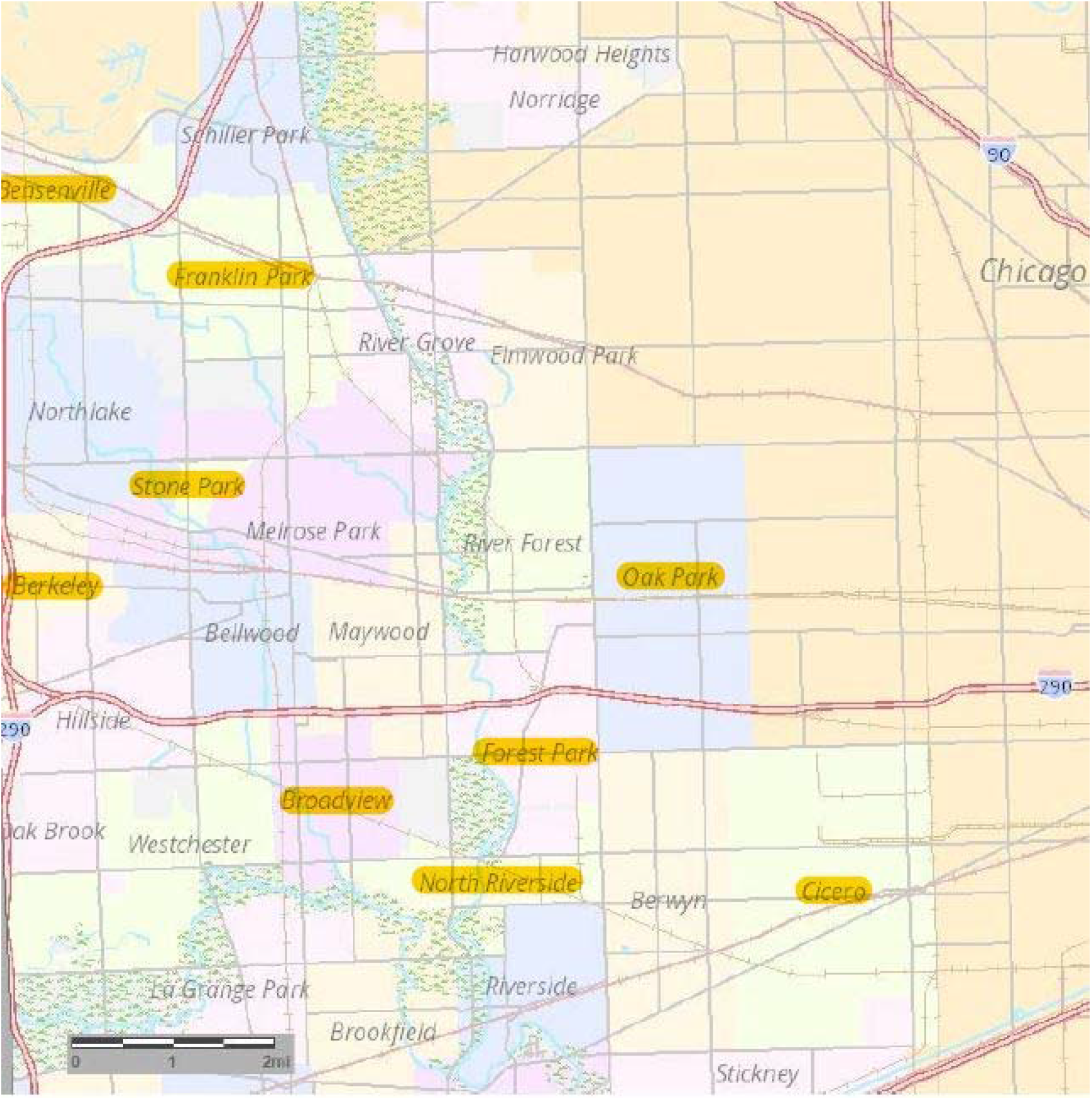
Highlighted final villages and townships with agreements Rush Mobile Stroke Unit at the time of pilot program closure

While our MSU was active and transporting patients, some difficulties arose. First, cellular signal or modem/router hardware encountered problems which occasionally led to dropped evaluations, and time out of operation to resolve issues. Second, delays in arrival of MSU when local EMS was ready to transport was encountered if calls were near coverage limit borders or recent/simultaneous call and transport. Third, mechanical problems with CT scanner and scanner bed were encountered if MSU was not level while obtaining imaging. Fourth, imaging at times was inadequate to allow for appropriate diagnosis. Fifth, there may have been delay or hesitancy in buy-in from local EMS services who already have close and established ties with their communities. Lastly, the discrepancy of number of thrombolysis in the ED versus in the MSU.

Our study does have some limitations. We did not have complete data on onset time to report on patient’s presenting within the golden hour of stroke treatment. We also do note the low number of patients treated in our setting, and even lower suspected LVO that underwent intervention, limiting analysis of this population. Outcome data was also incomplete due to the low number of patients treated. A strength of this study, however, is reporting of treatment times and experiences in a suburban setting, and limitations on local EMS coverage.

In conclusion, the Rush MSU demonstrated significant reduction of acute ischemic stroke treatment time with intravenous thrombolysis, but did not demonstrate the patient volume necessary to justify continued operation. Suburban and rural regions do benefit from pre-hospital stroke evaluation, however the ideal method for a cost-effective strategy is still unknown.

## Data Availability

All data available was reported in the manuscript.

## Acknowledgements

All authors were a part of the study design and concept, manuscript process, revision, and are aware of submission.

